# SARS-CoV-2 genomic monitoring in the São Paulo State unveils new sublineages of the AY.43 strain

**DOI:** 10.1101/2021.11.29.21266819

**Authors:** Alex Ranieri Jerônimo Lima, Gabriela Ribeiro, Vincent Louis Viala, Loyze Paola Oliveira de Lima, Antonio Jorge Martins, Claudia Renata dos Santos Barros, Elaine Cristina Marqueze, Jardelina de Souza Todao Bernardino, Debora Botequio Moretti, Evandra Strazza Rodrigues, Elaine Vieira Santos, Ricardo Augusto Brassaloti, Raquel de Lello Rocha Campos Cassano, Pilar Drummond Sampaio Corrêa Mariani, Luan Gaspar Clemente, Patricia Akemi Assato, Felipe Allan da Silva da Costa, Mirele Daiana Poleti, Jessika Cristina Chagas Lesbon, Elisangela Chicaroni Mattos, Cecilia Artico Banho, Lívia Sacchetto, Marília Mazzi Moraes, Melissa Palmieri, Fabiana Erica Vilanova da Silva, Rejane Maria Tommasini Grotto, Jayme A. Souza-Neto, Marta Giovanetti, Luiz Carlos Junior de Alcantara, Maurício Lacerda Nogueira, Heidge Fukumasu, Luiz Lehmann Coutinho, Simone Kashima, Raul Machado Neto, Dimas Tadeu Covas, Svetoslav Nanev Slavov, Maria Carolina Elias, Sandra Coccuzzo Sampaio

## Abstract

Delta VOC is highly diverse and more than 120 sublineages have been identified in Pango lineages with the continuous description of emerging ones. Brazil is now one of the most vaccinated countries against SARS-CoV-2 in the world which can enhance the emergence of viral mutations related to improved viral fitness. In this study, we identified two novel sublineages of the AY.43 lineage which were classified as AY.43.1 and AY.43.2 as observed on the specific clustering on the obtained phylogenetic tree. The novel sublineages were defined by the following characteristic nonsynonymous mutations ORF1ab:A4133V and ORF3a:T14I for AY.43.1 and ORF1ab:G1155C for AY.43.2. The majority of the analyzed sequences of both lineages were Brazilian, which shows that probably these two emerging sublineages have Brazilian origin. It is still unknown how these two sublineages are disseminated in São Paulo State and Brazil and their potential impact on the ongoing vaccination process. However, the performed study reinforces the importance of the SARS-CoV-2 genome monitoring for timely identification of emerging SARS-CoV-2 variants which can impact the ongoing SARS-CoV-2 vaccination and public health policies.

## Introduction

Currently in Brazil have been applied more than 157 million doses of anti-SARS-CoV-2 vaccine and the individuals who have been fully vaccinated are approximately 124 million, which makes Brazil one of the most vaccinated nations in the world. In Brazil, on the background of this solid process of vaccination, the first cases of Delta VOC introductions were reported (1). Although now there is a full substitution of the dominant Brazilian gamma VOC by the Delta sublineages. Despite the full substitution of the pre-existing gamma VOC in Brazil with the delta VOC, no exponential growth of the new cases has been observed, most probably due to the ongoing vaccination. Delta VOC is highly diversified, and more than 120 sublineages have been classified within the Pango lineages (https://cov-lineages.org/) with the frequent description of novel lineages and sublineages.

We assume that the presence of massive vaccination in Brazil may also help for the selection of novel SARS-CoV-2 lineages and sublineages due to the accumulation of viral mutations leading to viral fitness. The molecular surveillance of the SARS-CoV-2 variants is of crucial importance for tracking the genomic profile of this virus and the accumulation of mutations which on one hand can alter the viral functions in terms of infectivity and transmissibility and on the other to be important for the emergence of novel variants, lineages, and sublineages which can exert significant pressure on the healthcare system of a given country (2).

In this study, we identified two novel Brazilian sublineages belonging to the AY.43 lineage, which were named AY.43.1 and AY.43.2, suggested due to their specific clade clustering within the main AY.43 lineage. Most of these sequences originated from the city of São Paulo and the genome monitoring was performed by the Network for Pandemic Alert of SARS-CoV-2 Variants of the São Paulo State.

## Materials and methods

The raw sequence data obtained were submitted to quality control analysis using the FastaQC (3) software version 0.11.8. Trimming was performed using Trimmomatic (4) version 0.3.9 in order to select the sequences with the best quality. Only sequences with quality scores >30 were used. We mapped the trimmed sequences against the SARS-CoV-2 reference (Genbank RefSeq NC_045512.2) using BWA (5) software and samtools for read indexing. The mapped files were submitted to refinement using Pilon (7) to obtain the correct indels and insertions. The trimmed sequences were subjected to a remap against the genome refined by Pilon. Finally, we use bcftools (8) for variant calling and seqtk (9) for the assembly of the consensus SARS-CoV-2 genomes. Positions covered by fewer than 10 reads (DP<10) and bases of quality lower than 30 were considered a gap in coverage and converted to Ns. Coverage values for each genome were calculated using samtools v1.12. We assessed the consensus genome sequence quality using Nextclade v0.8.1 (https://clades.nextstrain.org).

Phylogenetic analysis was performed using the Nextstrain v3.0.3 SARS-CoV-2 workflow (10). To reconstruct the phylogeny of the AY.43 hypothetical sublineages we used 1,616 SARS-CoV-2 sequences generated from the Butantan Network for Pandemic Alert of SARS-CoV-2 Variants distributed between the epidemiological weeks 41 to 43 (GISAID accession ID in Supplementary File 1). The analyzed sequences were obtained from all 17 Health Divisions of the São Paulo State. A representative global dataset was retrieved from GISAID (3.711 sequences, downloaded from nextregions global in 2021, 10th of November - GISAID accession ID in Supplementary File 2) (https://www.gisaid.org/) which was used as background for our analysis.

## Results

In the performed phylogenetic analysis we observed a large cluster of the AY.43 lineage containing 492 sequences, the majority of which (n=359, 73%) were from Brazil (Figure 1). From the Brazilian sequences, only one sequence was not generated from the Butantan Network for Pandemic Alert of SARS-CoV-2 Variants. An interactive tree can be accessed at the following link: https://nextstrain.org/fetch/repositorio.butantan.gov.br/bitstream/butantan/3990/1/ncov_EpiWeek_forty-one-to-forty-three.json. Additionally, the dissemination of these sequences in the São Paulo State showed that the majority were obtained from the metropolitan zone of São Paulo city.

**Figure 1.**
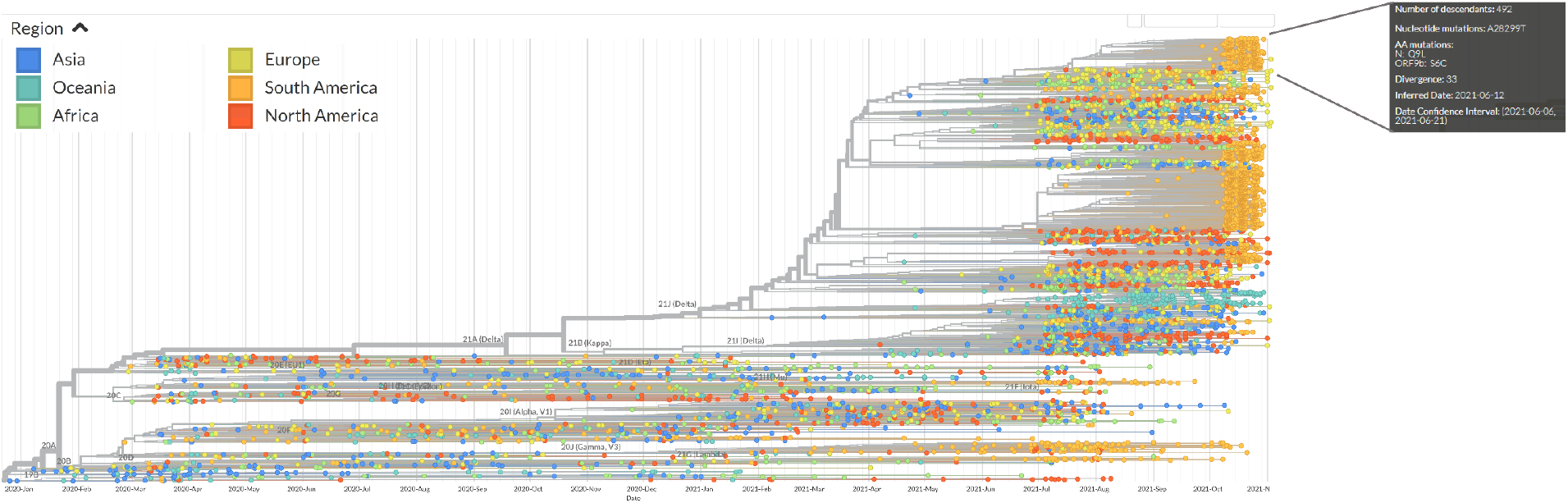
Nextstrain tree colored by continent. The AY.43 lineage clade is indicated by the lines, and the box contain its information, such as number of sequences in the clade and the amino acid mutations present in all sequences.

The Brazilian AY.43 sequences were divided as two subclades within the AY.43 cluster: (i) 104 sequences sharing the amino acid mutations in ORF1ab: A4133V and in ORF3a:T14I; named AY.43.1 (ii) and 99 sequences, which presented the amino acid mutation in ORF1ab:G1155C and named AY.43.2 (Figure 2). We additionally observed other subclusters in the AY.43 clade, including in the proposed AY.43.1 and AY.43.2 clades. However, these were composed of a limited number of sequences and did not show sufficient support to be suggested as novel sublineages of AY.43.

**Figure 2.**
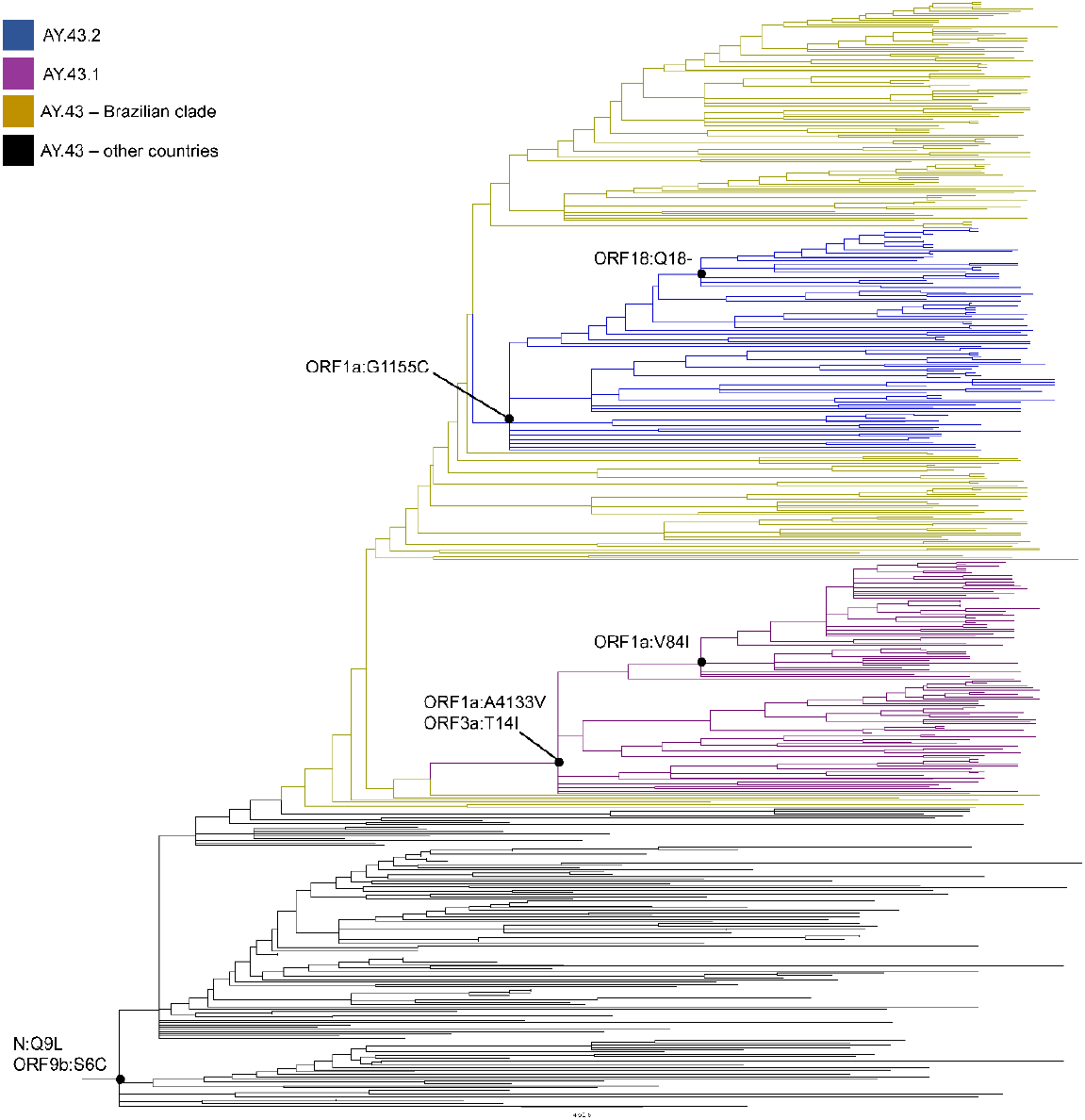
AY.43 clade zoomed from Figure 1. The yellow clade contains 359 Brazilian sequences, which form two subclades: AY.43.1 in purple and AY.43.2 in blue. Dots in branches indicates amino acid mutations present in all sequences from that clade.

## Discussion

In this study, we provide a preliminary report of the characterization of two novel AY.43 sublineages with probable Brazilian origin, due to the observed specific clustering and specific mutation profile observed in the phylogenetic tree generated by Nextstrain. Additionally, depending on the type of mutation (localization in ORF1a or ORF3a) we hypothesized the subdivision of the AY.43 strain into two sublineages designated as AY.43.1 and AY.43.2. The obtained data shows the importance of the SARS-CoV-2 genomic surveillance for the identification of emerging lineages. This is particularly important because SARS-CoV-2 emerging lineages can exert an enormous impact on the public health systems due to increased infectivity and transmission (2). In the newly characterized sublineages, we defined the mutations A4133V and G1155C in the ORF1a and the T14I mutation in the ORF3a, which were non-synonymous. To our knowledge, the impact of these mutations is unknown, except for the T14I, which shows deleterious effects on the viral proteins (11). A nonsynonymous mutation in ORF3a, which is a conserved protein involved in viral replication and release (12), may affect viral functions in addition to the mutational constellation defined for the Delta VOC. Based on the performed analysis the newly classified AY.43.1 and AY.43.2 sublineages probably have Brazilian origin as were composed of sequences obtained from Brazil. Further studies are necessary to investigate the dissemination in these two sublineages in Brazil, but preliminary results show that the majority of AY.43.1 and AY.43.2 sequences originated from the city of São Paulo.

In conclusion, we show that SARS-CoV-2 genome monitoring is crucial for the prompt characterization of SARS-CoV-2 novel lineages and sublineages. By this approach, we can timely detect the presence of novel SARS-CoV-2 variants and implement strategies for preventing their dissemination which can have further implications on the ongoing SARS-CoV-2 vaccination and public health policies.

## Data Availability

Genomes used in this study can be obtained from GISAID, using accession ID present in Supplementary Files 1 and 2.

https://repositorio.butantan.gov.br/handle/butantan/3990?locale=en

## Acknowledgements

We thank all contributors from GISAID. We also thank Gabriela Ribeiro and Glaucia Borges for their help with the Instituto Butantan’s local database repository.

## Funding

This work was supported by Butantan Foundation, Fundação de Amparo à Pesquisa do Estado de São Paulo (Grant Numbers: 2020/10127; 2020/06441-2), Fundação Hemocentro Ribeirão Preto

### Institutional Review Board Statement

The study was conducted according to the guidelines of the Declaration of Helsinki and approved by the Institutional Ethics Committee of the Faculty of Medicine of Ribeirão Preto, University of Sao Paulo (CAAE: 50367721.7.1001.5440).

### Informed Consent Statement

Patient consent was waived once the diagnostic test had already been performed and the result has been communicated to the patients. Viral RNA was used, and the results obtained will not compromise private information, neither bring any new clinical/therapeutic outcome for the patient.

